# Inequalities in cancer diagnostic outcomes for patients with a learning disability: a retrospective cohort study in England

**DOI:** 10.64898/2026.02.03.26345112

**Authors:** B Wiering, GA Abel, L Farmer, R Kerrison, SWD Merriel, SJ Price, D Shotter, JM Valderas, LTA Mounce

**Affiliations:** Department of Health and Community Sciences, Faculty of Health and Life Sciences, University of Exeter, St Luke’s Campus, Heavitree Road, Exeter, EX1 2LU United Kingdom; Cancer Care Expert Group, School of Health Sciences, University of Surrey, Guildford, Surrey, GU2 7XH United Kingdom; Centre for Primary Care & Health Services Research, School of Health Sciences, Faculty of Biology, Medicine and Health, The University of Manchester, Oxford Rd, Manchester, M13 9PL United Kingdom; Department of Family Medicine & Centre for Research on Health Systems Performance of the National University Health System, National University of Singapore, 21 Lower Kent Ridge Road, 119077 Singapore

**Keywords:** Learning disability, Intellectual disability, Cancer stage, Mortality

## Abstract

**Objectives:** The cancer diagnostic process may be more complicated for patients with a learning disability (“intellectual disability” outside of the UK) than for other patients. We aimed to investigate whether patients with a learning disability were more likely to experience disadvantage in cancer diagnostic pathways and outcomes.

**Design:** A retrospective cohort study using routinely collected linked data across primary and secondary care, and cancer registration.

**Setting:** 1470 general practices in England.

**Participants:** 277,050 participants who were aged 40+ years, had an incident cancer recorded in the cancer registration during 2012-2018 and were registered for at least three years at their general practice prior to diagnosis.

**Main outcome measures:** Emergency presentation route to diagnosis or urgent suspected cancer referral route to diagnosis, cancer stage at diagnosis (early vs advanced) and all-cause mortality within 30 days after diagnosis.

**Results:** 277,050 patients were included in the study, with 796 (0.3%) patients having a record of a learning disability. Patients with a learning disability were over twice as likely to be diagnosed via an emergency presentation (aOR:2.65, 95%CI:2.26-3.11, p<0.001), and half as likely to be diagnosed via an urgent suspected cancer referral (aOR:0.51, 95%CI:0.43-0.60, p<0.001). They were also more likely to be diagnosed with advanced-stage cancer (aOR:1.37, 95%CI:1.12-1.67, p=0.002), especially for breast cancer, and to die within 30 days of diagnosis (aOR:3.77, 95%CI:3.10-4.59, p<0.001) than patients without a learning disability.

**Conclusions:** Patients with a learning disability experience marked inequalities in cancer diagnostic pathways and outcomes. The increased risk of advanced-stage breast cancer is of particular note. Improved support to access and navigate the health care system may be required to negate experienced difficulties during the diagnostic process.

**Summary box:** *What is already known on this topic:*

- Patients with learning disabilities experience avoidable mortality after a cancer diagnosis.
- Greater dependence on others, low symptom awareness and difficulties accessing and navigating the health care system during the cancer diagnostic process potentially contribute to worse outcomes, particularly for more severe learning disability.

*What this study adds:*

- Considering all cancer sites except non-melanoma skin cancer, we found that having a record of a learning disability was associated with an increased likelihood of being diagnosed at an advanced stage, especially for breast cancer, and increased mortality within 30 days after diagnosis.
- Patients with a learning disability were more likely to be diagnosed as an emergency, and less likely to be diagnosed via an urgent suspected cancer referral.

*How this study might affect research, practice or policy:*

- Patients with a learning disability require better support to negate existing inequalities in diagnostic pathways and outcomes

## Introduction

Cancer survival rates have improved greatly in recent decades.[1-3] Some patient groups, however, such as patients with a learning disability – synonymous with “intellectual disability”, as preferred outside of the UK,[4] still experience significant avoidable cancer mortality.[5] A learning disability is defined as a lifelong impaired intellectual ability, and reduced social functioning. It covers varying levels of intellectual ability, experienced difficulties and support needs, and is estimated to affect around 1-3% of the population worldwide.[6] Improving care for patients with a learning disability is a priority for the English National Health Service (NHS), and is included in the NHS long term plan,[7] the Health and Social Care Act 2022,[8] and the Core20PLUS5 approach to tackling healthcare inequalities.[9] These efforts are very timely, as an English report on the lives and deaths of patients with a learning disability found that cancer was the second most common cause of death and accounted for 15.7% of avoidable deaths.[10]

A potential reason for avoidable cancer mortality may be found in delayed diagnosis. Patient outcomes tend to be better if patients are diagnosed early, as it increases the number of available treatment options and their success, while reducing treatment length, complications, and long-term cancer- and treatment impacts.[11, 12] People with learning disabilities, however, experience difficulties in recognising possible cancer symptoms,[13] communicating their experiences, and accessing and navigating the health care system.[14-16] A recent review suggested that people with learning disabilities, their carers, and health care practitioners have low awareness of possible cancer symptoms, and that carers and health care professionals often were not sure of their role in facilitating cancer awareness for patients with a learning disability.[13] Ethnographic studies found that people with a learning disability may have higher levels of dependence on others to recognise, acknowledge and explain possible cancer symptoms, and help them access and navigate the health care system. Additionally, carers often have an important role in what information is shared, and in deciding whether a person with a learning disability is able to cope with the cancer diagnostic process and subsequent treatments.[15, 17]

These factors are likely to complicate the diagnostic process, and could contribute to a delayed diagnosis.[15] Our NIHR-funded programme (Spotting Cancer among Comorbidities (SPOCC), NIHR201070) explored cancer diagnostic processes and outcomes in patients with 121 pre-existing health conditions. Here, we present findings relating to the following research questions:

1. Are patients with a learning disability more likely to experience suboptimal pathways to diagnosis than patients without learning disabilities (i.e. increased emergency presentations and decreased urgent suspected cancer referrals in primary care)?
2. Are patients with a learning disability more likely to be diagnosed with advanced-stage cancer than patients without a learning disability?
3. Are patients with a learning disability more likely to die within 30 days of their cancer diagnosis than patients without a learning disability?

## Materials and methods

### Data source

The Clinical Practice Research Datalink (CPRD) is an electronic database of routinely collected de-identified primary care health records in the UK. For this study, CPRD Aurum data relating to English patients was used. CPRD Aurum covered 10% of GP practices in England in 2018.[18] This data was linked to the National Cancer Registration and Analysis Service (NCRAS),[19] the patient-level Index of Multiple Deprivation (IMD) 2015 and National Office for Statistics death data. Linkage was carried out by a trusted third party at NHS England. No patient-identifiable data is received by researchers using the linked data.[20] CPRD linked datasets are validated datasets, which have widely been used for cancer diagnostic studies.[21-25]

### Study population

We included patients aged 40 years and older, with primary care data available in CPRD, an incident cancer diagnosis recorded in NCRAS data between 01/01/2012 and 31/12/2018, and three years continued registration at their practice before their cancer diagnosis. Patients were excluded if they: 1) had a recorded sex not matching a sex-specific cancer (e.g. male patients with uterine cancer); 2) had a cancer diagnosis recorded before the inclusion time-period; 3) were diagnosed via screening; 4) were diagnosed with non-melanoma skin cancer; 5) died before their diagnosis.

### Ethical approval

Ethical approval has been granted for observational research using anonymised CPRD data by the National Research Ethics Service (NRES). Study-specific approval is therefore not required.[26] Protocol approval was granted by the CPRD Independent Scientific Advisory Committee (ISAC – Protocol 20_126R).

### Patient and public involvement (PPI)

The study is embedded in a Programme Grant, which was designed with extensive PPI consultation. The programme has a patient co-investigator, and a PPI group, with PPI members fulfilling bridge roles for work packages.[27] For this study, the PPI bridge (LF) attended three work package meetings and contributed to the manuscript. One meeting was held with the PPI group to disseminate results.

### Patient characteristics

Patient age, sex (male/female), and smoking history (never/ever) were extracted from CPRD. Age was treated as a categorical variable with 5-year age groups between 40 and 89 years and a 90 years and older group. IMD groups were defined based on national quintiles. The Cambridge Multimorbidity Score (CMS) general outcome weighting[28] was used to measure morbidity burden at least a year before diagnosis. The CMS is a validated measure consisting of 37 long-term conditions, and is weighted by primary care use, mortality and unplanned hospital admissions.[28] Where available, pre-existing validated code lists were used to capture CMS conditions. New code lists were developed using robust methods.[29] Four morbidity burden groups (none, low, medium, high) were created, with the last three based on CMS tertiles excluding the weighting for learning disability.

### Learning disability

Learning disability was measured using definitions developed for the CMS.[28] A validated code list developed for the Quality and Outcomes Framework (business rules version 45) was used.[30] The Quality and Outcomes Framework is a pay-for-performance scheme in the UK that rewards GP practices based on indicators, one of which is to maintain a register of patients with a learning disability.[31] Patients were categorised as having a learning disability if a learning disability code was recorded at least 12 months before their cancer diagnosis.

### Cancer site and stage

Cancer diagnoses and stage were extracted from NCRAS, and the tumour with the earliest diagnosis date was included. In rare cases where multiple diagnoses were recorded on the same date, the most advanced-stage cancer was retained. A random selection was made for any remaining multiple diagnoses such that each patient had only a single tumour included in the analysis. Based on ICD-10 codes, 25 common sites plus ductal carcinoma in situ (DCIS) were identified with remaining cancers assigned to an “other” category (codes C00-C97 and D05.1, but not C44; Appendix A, Table 1).

**Table 1.** Patient characteristics for patients with and without a learning disability

|  | No learning disability<br>(n=276254) | Learning disability (n=796) | Total (n=277050) |
| --- | --- | --- | --- |
|  | N (%) | N (%) | N (%) |
| *Age* |  |  |  |
| 40 to 44 years | 7117<br>(2.6%) | 47<br>(5.9%) | 7164<br>(2.6%) |
| 45 to 49 years | 12686<br>(4.6%) | 67<br>(8.4%) | 12753<br>(4.6%) |
| 50 to 54 years | 17048<br>(6.2%) | 94<br>(11.8%) | 17142<br>(6.2%) |
| 55 to 59 years | 22999<br>(8.3%) | 119<br>(15.0%) | 23118<br>(8.3%) |
| 60 to 64 years | 29783<br>(10.8%) | 131<br>(16.5%) | 29914<br>(10.8%) |
| 65 to 69 years | 40268<br>(14.6%) | 111<br>(13.9%) | 40379<br>(14.6%) |
| 70 to 74 years | 41974<br>(15.2%) | 102<br>(12.8%) | 42076<br>(15.2%) |
| 75 to 79 years | 39403<br>(14.3%) | 69<br>(8.7%) | 39472<br>(14.3%) |
| 80 to 84 years | 32610<br>(11.8%) | 33<br>(4.2%) | 32643<br>(11.8%) |
| 85 to 89 years | 21040<br>(7.6%) | 20<br>(2.5%) | 21060<br>(7.6%) |
| 90 years and older | 11326<br>(4.1%) | 3<br>(0.4%) | 11329<br>(4.1%) |
| Sex (Female) | 128106<br>(46.4%) | 360<br>(45.2%) | 128466<br>(46.4%) |
| *IMD* |  |  |  |
| 1 (least deprived) | 63928<br>(23.2%) | 93<br>(11.7%) | 64021<br>(23.1%) |
| 2 | 59384<br>(21.5%) | 128<br>(16.1%) | 59512<br>(21.5%) |
| 3 | 54112<br>(19.6%) | 188<br>(23.6%) | 54300<br>(19.6%) |
| 4 | 50871<br>(18.4%) | 174<br>(21.9%) | 51045<br>(18.4%) |
| 5 (most deprived) | 47878<br>(17.3%) | 213<br>(26.8%) | 48091<br>(17.4%) |
| *Ethnicity* |  |  |  |
| White | 256125<br>(92.7%) | 755<br>(94.9%) | 256880<br>(92.7%) |
| Asian | 7492<br>(2.7%) | 14<br>(1.8%) | 7506<br>(2.7%) |
| Black | 6605<br>(2.4%) | 14<br>(1.8%) | 6619<br>(2.4%) |
| Other | 1564<br>(0.6%) | 0<br>(0.0%) | 1564<br>(0.6%) |
| Mixed | 1408<br>(0.5%) | 7<br>(0.9%) | 1415<br>(0.5%) |
| Unknown | 3060<br>(1.1%) | 6<br>(0.8%) | 3066<br>(1.1%) |
| *Morbidity burden* |  |  |  |
| No morbidity burden | 43951<br>(15.9%) | 66<br>(8.3%) | 44017<br>(15.9%) |
| Low morbidity burden | 77934<br>(28.2%) | 124<br>(15.6%) | 78058<br>(28.2%) |
| Medium morbidity burden | 77321<br>(28.0%) | 299<br>(37.6%) | 77620<br>(28.0%) |
| High morbidity burden | 77048<br>(27.9%) | 307<br>(38.6%) | 77355<br>(27.9%) |
| *Cancer type* |  |  |  |
| Bladder | 7748<br>(2.8%) | 21<br>(2.6%) | 7769<br>(2.8%) |
| Brain | 3855<br>(1.4%) | 15<br>(1.9%) | 3870<br>(1.4%) |
| Breast | 29753<br>(10.8%) | 77<br>(9.7%) | 29830<br>(10.8%) |
| Cervix | 1266<br>(0.5%) | 2<br>(0.3%) | 1268<br>(0.5%) |
| Colon | 21330<br>(7.7%) | 94<br>(11.8%) | 21424<br>(7.7%) |
| DCIS | 2622<br>(1.0%) | 6<br>(0.8%) | 2628<br>(1.0%) |
| HL | 1025<br>(0.4%) | 2<br>(0.3%) | 1027<br>(0.4%) |
| Larynx | 1731<br>(0.6%) | 2<br>(0.3%) | 1733<br>(0.6%) |
| Leukaemia | 7071<br>(2.6%) | 23<br>(2.9%) | 7094<br>(2.6%) |
| Liver | 4919<br>(1.8%) | 20<br>(2.5%) | 4939<br>(1.8%) |
| Lung | 36687<br>(13.3%) | 62<br>(7.8%) | 36749<br>(13.3%) |
| Melanoma | 11060<br>(4.0%) | 22<br>(2.8%) | 11082<br>(4.0%) |
| Mesothelioma | 2261<br>(0.8%) | 4<br>(0.5%) | 2265<br>(0.8%) |
| Myeloma | 4907<br>(1.8%) | 10<br>(1.3%) | 4917<br>(1.8%) |
| NHL | 10965<br>(4.0%) | 34<br>(4.3%) | 10999<br>(4.0%) |
| Oesophagus | 7780<br>(2.8%) | 36<br>(4.5%) | 7816<br>(2.8%) |
| Oral | 7061<br>(2.6%) | 10<br>(1.3%) | 7071<br>(2.6%) |
| Other | 18544<br>(6.7%) | 101<br>(12.7%) | 18645<br>(6.7%) |
| Ovary | 6111<br>(2.2%) | 29<br>(3.6%) | 6140<br>(2.2%) |
| Pancreas | 8566 | 18 | 8584 |
|  | (3.1%) | (2.3%) | (3.1%) |
| Prostate | 44830 | 57 | 44887 |
|  | (16.2%) | (7.2%) | (16.2%) |
| Rectum | 10938 | 48 | 10986 |
|  | (4.0%) | (6.0%) | (4.0%) |
| Renal | 8669 | 28 | 8697 |
|  | (3.1%) | (3.5%) | (3.1%) |
| Stomach | 5393 | 18 | 5411 |
|  | (2.0%) | (2.3%) | (2.0%) |
| Testis | 895 | 7 | 902 |
|  | (0.3%) | (0.9%) | (0.3%) |
| Thyroid | 2304 | 3 | 2307 |
|  | (0.8%) | (0.4%) | (0.8%) |
| Uterus | 7963 | 47 | 8010 |
|  | (2.9%) | (5.9%) | (2.9%) |

Cancer stage was divided into two groups; early stage included cancer stages 0, I and II, and advanced stage included stages III and IV. Brain cancer and leukaemia are not stageable in the system used for staging (TNM), and were therefore excluded from stage analyses (Please see Appendix A, Table 2 for missing stage information).

**Table 2.**
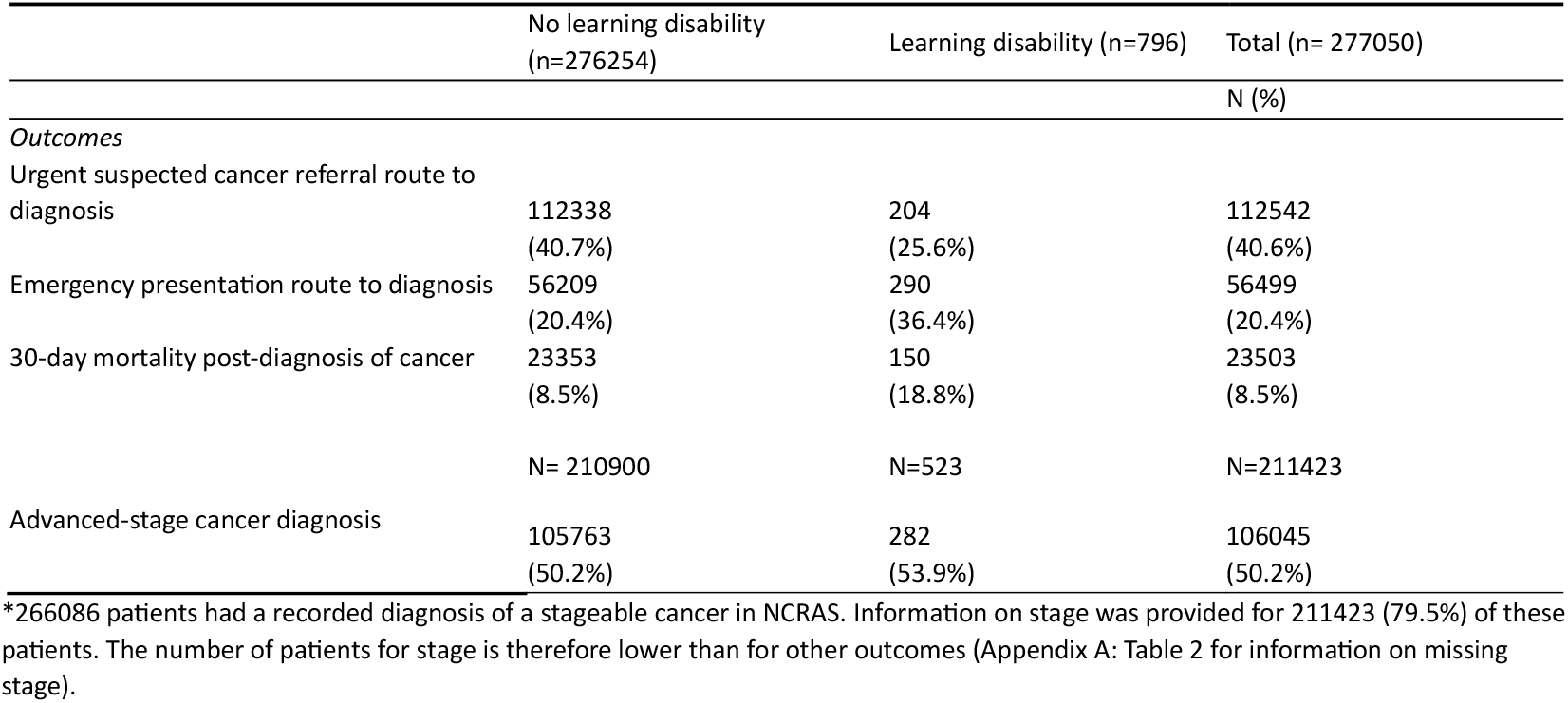
Outcomes and routes to diagnosis for patients with and without a learning disability

### Mortality and route to diagnosis

Office for National Statistics death data was used to establish date of death. Based on this date, a binary variable was created that indicated whether a patient had died of any cause within 30 days after their cancer diagnosis. Route to diagnosis data in NCRAS was used to extract pathways leading directly to diagnosis. Examples of these pathways are diagnoses after an urgent suspected cancer referral or emergency presentation.[32] An emergency presentation includes emergency admission, referral, transfer, or attendance to- or within secondary care. Two binary variables were created, one indicating whether patients were diagnosed via an emergency presentation (vs any other route), and the other indicating whether patients were diagnosed via urgent suspected cancer referral.

### Data analysis

Logistic regression models were used to investigate whether patients with a learning disability were more likely to: be diagnosed at advanced stage, be diagnosed via an emergency route, be diagnosed via an urgent suspected cancer referral, or die within 30 days of diagnosis. Initial main effects-only models included patient age, sex, cancer site, an indicator variable for learning disability, deprivation quintile, morbidity burden, smoking history and year of diagnosis. Clustering of observations at general practice-level was accounted for by a standard error adjustment. In order to investigate cancer site specific differences, two-way interaction terms between cancer site and patient characteristics, and cancer site and the presence of a learning disability were investigated individually with all interactions retained in the final model. A joint Wald test was used to test for significance. For most variables included in the models, completeness of records was assumed, with the absence of a record indicating the absence of a diagnosis. Missing data on deprivation (0.03%) and cancer stage (20.5%) was considered missing at random. Analyses with missing outcome data are unbiased under the Missing at Random assumption. Approaches such as multiple imputation were therefore not needed. Complete case analysis was conducted for every model. Analyses were conducted using Stata v.17[33].

## Results

We included 277,050 patients in the study, of which 796 (0.3%) patients had a record of a learning disability. Patients with a learning disability were generally younger than patients without a learning disability, with an average age of 63 (SD:11.3; Range:40.1-94.2) compared to 70.2 years (SD:12.4; Range:40-107.8). The most common cancer sites for patients with a learning disability were colon (n=94; 11.8%) and breast (n=77; 9.7%), while for patients without learning disabilities, the most common cancers were prostate (n=44,830; 16.2%) and lung (n= 36,687; 13.3%) (Table 1).

Patients with a learning disability had higher rates of emergency diagnoses (36.4% (n=290) compared to 20.4% (n=56,209)) and fewer diagnoses via an urgent suspected cancer referral than patients without a learning disability (25.6% (n=204) compared to 40.7% (n= 112,338)). Patients with a learning disability also had a higher number of advanced stage diagnoses (53.9% (n=282) compared to 50.2% (n= 105,763) (Appendix A, Table 2 for missing stage information) and higher 30-day mortality rates (18.8% (n= 150) compared to 8.5% (n= 23,353)) than patients without a learning disability (Table 2).

### Routes to diagnosis

After adjusting for age, sex, deprivation, morbidity burden, smoking history, cancer site and year of diagnosis, patients with a learning disability were much more likely to be diagnosed via emergency presentations (aOR:2.65, 95%CI:2.26-3.11, p<0.001), and much less likely via urgent suspected cancer referrals (aOR:0.51, 95%CI:0.43-0.60, p<0.001) than patients without a learning disability (Table 3; Appendix A, Table 3). The interaction effect model for emergency presentation found evidence (p<0.05) that patients with a learning disability were more likely to be diagnosed after an emergency presentation for the majority of cancer sites (Appendix A, Figure 1), while an interaction effects model for urgent suspected cancer referral found evidence (p<0.05) that patients with a learning disability who were diagnosed with oral, lung, ovarian, prostate, uterus, other, rectum, colon, testicular, and breast cancer were less likely to be diagnosed after an urgent suspected cancer referral than patients without a learning disability (Appendix A, Figure 2).

**Table 3.** Associations between emergency presentation route to diagnosis, urgent referral route to diagnosis, cancer stage at diagnosis, 30-day mortality and learning disability

|  | Learning disability (ref.: no learning disability) * |  |  |  |
| --- | --- | --- | --- | --- |
|  | Odds ratio | Lower 95% confidence interval | Upper 95% confidence interval | P |
| *Outcomes* |  |  |  |  |
| Urgent suspected cancer referral route to diagnosis | 0.51 | 0.43 | 0.60 | P<0.001 |
| Emergency presentation route to diagnosis | 2.65 | 2.26 | 3.11 | P<0.001 |
| 30-day mortality post-diagnosis of cancer | 3.77 | 3.10 | 4.59 | P<0.001 |
| Advanced-stage cancer diagnosis | 1.37 | 1.12 | 1.67 | 0.002 |

\*Separate regression models were run per outcome. Analyses were adjusted for patient age, sex, cancer site, an indicator variable for learning disability, deprivation quintile, morbidity burden, smoking history and year of diagnosis.

### Cancer stage and mortality

After adjusting for age, sex, deprivation, morbidity burden, smoking history, cancer site and year of diagnosis, patients with a learning disability were more likely to be diagnosed with advanced-stage cancer (aOR:1.37, 95%CI:1.12-1.67, p=0.002) and to die within 30 days after their diagnosis (aOR:3.77, 95%CI:3.10-4.59, p<0.001) than patients without a learning disability (Table 3; Appendix A, Table 4). Interaction effects models for cancer stage and mortality found evidence (p<0.05) that effects of learning disability vary by cancer site. For stage, patients with a learning disability were more likely to be diagnosed at advanced stage for breast (aOR:3.36, 95%CI:1.99-5.65, p<0.001) and ‘other’ cancers (aOR:3.87, 95%CI:1.34-11.17, p=0.012) (Appendix A, Figure 3). For mortality, patients with a learning disability diagnosed with all cancers apart from uterus, stomach, pancreatic and rectal cancer and non-Hodgkin lymphoma, were more likely to die within 30 days of their cancer diagnosis than patients without a learning disability (Appendix A, Figure 4). Large uncertainty due to limited numbers meant that many substantial point estimates for individual cancer sites did not reach statistical significance and it should be noted that for some sites point estimates were consistent with patients with a learning disability being diagnosed at earlier stages than patients without (oral, uterus, bladder, pancreas, mesothelioma, renal, stomach and melanoma) (Appendix A, Figure 3). For 30-day mortality, all point estimates favoured those without learning disabilities.

## Discussion

### Statement of principal findings

The present study investigated whether patients with a learning disability diagnosed with any cancer experience inequalities in diagnostic processes and outcomes. We found that patients with a learning disability experience significant disadvantages; having over twice the odds of being diagnosed via an emergency diagnosis and nearly half the odds via the preferred route of an urgent suspected cancer referral, compared with patients without learning disabilities. This study also found that patients with a learning disability are 37% more likely to be diagnosed at an advanced stage – over three times as likely for breast cancer – and have close to 4 times the odds of dying within 30 days after their cancer diagnosis.

### Comparison with existing literature

Previous research suggests that patients with a learning disability may experience difficulties in communicating their symptoms, and diagnostic overshadowing – where symptoms of a condition are erroneously attributed to their learning disability.[15, 16, 34] This could result in the cancer not being recognised by the carer, or in primary care, which potentially contributes to patients not being referred for cancer investigations and an increase in emergency diagnoses once the symptoms become urgent. To better understand whether possible cancer symptoms are not acted on in primary care, research is needed to investigate presenting patterns and resulting GP actions for patients with a learning disability. An alternative explanation for our findings is that the patient, carer and/or health care professional decide to not pursue cancer investigations,[15] and the diagnosis is made once the patient presents as an emergency. Even when adjusted for cancer stage, emergency diagnosis is commonly linked to worse patient outcomes.[32, 35] It is therefore important for future research to provide more insight into how patients with a learning disability come to be diagnosed as an emergency, and for carers and health care professionals to support cancer diagnosis through other routes.

The present study also found that patients with a learning disability are more likely to be diagnosed at an advanced stage, particularly if they are diagnosed with breast cancer, for which their odds are over threefold that of those without a learning disability. Existing evidence surrounding diagnostic outcomes is limited, but our findings are in line with a previous study suggesting that patients with learning disabilities are generally more likely to be diagnosed at an advanced stage[36, 37], and one small study suggesting that breast cancer patients with a learning disability were more likely to be diagnosed at an advanced stage than those without.[38] The finding regarding breast cancer is especially notable, as, although a greater likelihood of being diagnosed with advanced cancer could in part be explained by a lower uptake of screening,[39] a previous study found that for most patients breast cancer is generally quickly identified in primary care.[40] Future research should investigate whether associations of breast cancer symptoms with incident cancer are different for patients with and without a learning disability. In addition, improved support for patients with a learning disability to attend screening, and increased alertness to patients with a learning disability who present with possible breast cancer symptoms in primary care may help diagnose breast cancer earlier.

On top of an increased likelihood of being diagnosed with advanced-stage cancer, our results suggest that patients with a learning disability also experienced increased mortality shortly after being diagnosed with cancer. Although earlier research indicated that patients with a learning disability had greater cancer mortality rates than patients without a learning disability,[37, 41, 42] no studies have previously investigated short-term mortality. We do not yet know to what extent increased mortality is caused by; 1) patients with a learning disability being diagnosed at a point where cancer is an immediate threat to life; 2) cancer being diagnosed during emergency presentations for other life threatening conditions; 3) the presence of a learning disability affecting short-term care; 4), or a lower life expectancy for patients with a learning disability.[43] Our findings of suboptimal pathways to diagnosis and poor outcomes combined with previous findings regarding low cancer awareness, and difficulties navigating the diagnostic process,[15, 17] however, indicate a need for significant improvements in cancer awareness and diagnostic processes for patients with a learning disability.

### Strengths and weaknesses of the study

This is the first English retrospective cohort study using health care records to investigate diagnostic pathways and outcomes for patients with a learning disability diagnosed with any cancer. Some limitations, however, may have affected our findings. First, the work described here is a small part of a work package embedded in a Programme Grant. The work package aimed to explore the diagnosis of cancer in patients with pre-existing health conditions and studied 10 outcomes and 121 health conditions. The results for learning disability are discussed here, as this group experienced the worst outcomes of all patient groups, and the findings are likely to be of particular interest to researchers and policy makers. However, these findings should be considered in the light of the multiple analyses undertaken.

Only 0.3% of our study population had a record of a learning disability, while estimates suggest that approximately 2.5% of the English population has a learning disability.[15] This is in line with previous evidence suggesting that health records only re?ect up to a 0.5% prevalence of learning disabilities in the English population.[15, 44, 45] It is possible that recordings of learning disabilities were missed as they were recorded in ‘free/uncoded text’, which is not accessible to researchers. As there is generally a high overlap between free-text and diagnoses codes,[46] a more likely cause is that the code list only includes disorders where the prevalence of having a learning disability is 80 percent or higher.[30] Thus, it may be that we did not capture many cases with milder learning disability. We were not able to take into account the severity of the learning disability, or the presence of other complicating factors such as presence of autism or severe mental illness, which are more common in people with a learning disability.[47, 48]

Though we captured data on all cases in the cancer registry data with linked primary care data over a 7-year period, small sample sizes for individual cancer sites, and in particular for less common sites, meant that many estimates did not reach statistical significance. The absence of statistically significant associations should not be treated as evidence that such an association does not exist.

## Conclusion

Patients with a learning disability experience marked inequalities in cancer diagnostic pathways and outcomes. They are more likely to experience unfavourable routes to diagnosis, increased short-term mortality and are more likely to be diagnosed at an advanced stage than patients without learning disabilities. Previous evidence suggested that patients may have low symptom awareness and experience difficulties accessing and navigating care, which may have contributed to the inequalities highlighted by this study. Greater efforts need to be made to improve cancer awareness and support patients with a learning disability to access and navigate the health care system to improve cancer diagnostic outcomes.

## Statements

### Contributorship statement

The study was conceived by GA and LM, building on previous work by LM. BW curated and analysed the data supported by DS, LM and SP and produced the first draft of the paper with supervision of GA and LM. BW and LM directly accessed and verified the underlying data reported in the manuscript. All authors had access to the data, interpreted findings and identified issues for discussion. The draft was reviewed and modified with input from all authors over a number of versions. All authors saw and approved the final version. The corresponding author attests that all listed authors meet authorship criteria and that no others meeting the criteria have been omitted. Gary Abel is guarantor.

## Acknowledgements

This study is based in part on data from the Clinical Practice Research Datalink obtained under licence from the UK Medicines and Healthcare products Regulatory Agency. The data is provided by patients and collected by the NHS as part of their care and support. The interpretation and conclusions contained in this study are those of the author/s alone. The study is funded by the NIHR Programme Grants for Applied Research (PGfAR) SPOtting Cancer among Comorbidities (SPOCC) programme: supporting clinical decision making in patients with symptoms of cancer and pre-existing conditions (NIHR201070). SWDM is supported by the NIHR Manchester Biomedical Research Centre (NIHR203308). RK is supported by a Cancer Research UK Career Establishment Award (RCCCEA-Nov24/100001) and is a co-investigator on the NIHR Policy Research Programme Unit on Cancer Awareness, Screening and Early Diagnosis (reference PR-PRU-NIHR206132)). The views expressed are those of the authors and not necessarily those of the University of Surrey, NIHR or the Department of Health and Social Care.

## Data Availability Statement

Restrictions apply to the availability of these data. Routinely collected patient electronic health data was provided by CPRD. Access to data from CPRD is subject to a licence agreement and protocol approval.

## Declaration of generative AI in scientific writing

Generative AI and AI-assisted technologies were not used in the preparation of this work.

## Ethics approval

The National Research Ethics Service (NRES) has granted ethical approval for observational research using anonymised CPRD data. Observational studies using anonymised CPRD data are therefore not required to obtain study-specific ethical approval. Study protocol approval was granted by the CPRD Independent Scientific Advisory Committee (protocol 20_126R).

## Consent to participate

Patient consent was waived due to the nature of the data collection. GP practices choose to share patient data with CPRD for public health research purposes. No information that can identify a patient is ever sent to CPRD from the contributing GP practices. Because patients can’t be identified from data a GP practice sends to CPRD, the GP practice does not need to seek a patient’s consent to share data with CPRD. Individual patients can opt-out of sharing their data for research. CPRD does not collect data for these patients.

## Consent to publish

No identifiable patient data was made available to the research team by CPRD. Consent for publication has therefore not been sought.

## Transparency statement

The manuscript is an honest, accurate, and transparent account of the study being reported; no important aspects of the study have been omitted and there are no discrepancies from the study as originally planned.

