## Appendix A for "Inequalities in cancer diagnostic outcomes for patients with a learning disability: a retrospective cohort study in England"

**Table 1. ICD-10 codes for each cancer site included in the study**

| Cancer site | ICD-10 code |
|---|---|
| Lip, oral cavity and pharynx | C00, C01, C02, C03, C04, C05, C06, C07, C08, C09, C10, C11, C12, C13, C14 |
| Oesophagus | C15 |
| Liver | C22 |
| Stomach | C16 |
| Colon | C18 |
| Rectum | C19, C20 |
| Pancreas | C25 |
| Larynx | C32 |
| Lung | C33, C34 |
| Melanoma | C43 |
| Mesothelioma | C45 |
| Breast | C50 |
| Cervix | C53 |
| Uterus | C54, C55 |
| Ovary | C56, C57 |
| Prostate | C61 |
| Testis | C62 |
| Kidney | C64 |
| Bladder | C67 |
| Brain | C71 |
| Thyroid | C73 |
| Hodgkin lymphoma | C81 |
| Non-Hodgkin lymphoma | C82, C83, C84, C85 |
| Myeloma | C90 |
| Leukaemia | C91, C92, C93, C94, C95 |
| DCIS | D05 |

**Table 2. Missing cancer stage for stageable cancers for patients with and without a learning disability**

| | **Stageable cancers** | **Missing cancer stage for stageable cancers** |
|---|---|---|
| | N (%) | N (%)* |
| No learning disability | 265328 (99.7%) | 54428 (20.5%) |
| Learning disability | 758 (0.3%) | 235 (31.0%) |
| Total | 266086 (100%) | 54663 (20.5%) |

*Percentages presented in this column are row percentages and indicate the percentage of missing stage records for stageable cancers per group.

**Table 3. Main effect only model associations between emergency presentation route to diagnosis, urgent (two-week-wait) referral route to diagnosis, patient characteristics, cancer type and learning disability**

| | **Emergency presentation** | | | | **Urgent suspected cancer referral** | | | |
|---|---|---|---|---|---|---|---|---|
| | Odds ratio | Lower 95% confidence interval | Upper 95% confidence interval | P | Odds ratio | Lower 95% confidence interval | Upper 95% confidence interval | P |
| *Cancer type* | | | | P<0.001 | | | | P<0.001 |
| Bladder | 0.42 | 0.39 | 0.45 | | 2.16 | 2.04 | 2.28 | |
| Brain | 3.49 | 3.23 | 3.77 | | 0.03 | 0.03 | 0.04 | |
| Breast | 0.14 | 0.14 | 0.15 | | 7.55 | 7.20 | 7.92 | |
| Cervix | 0.55 | 0.47 | 0.65 | | 1.38 | 1.22 | 1.56 | |
| Colon | 1.00 | 1.00 | 1.00 | | 1.00 | 1.00 | 1.00 | |
| DCIS | 0.08 | 0.06 | 0.10 | | 1.74 | 1.60 | 1.90 | |
| HL | 0.55 | 0.46 | 0.66 | | 1.36 | 1.20 | 1.55 | |
| Larynx | 0.28 | 0.24 | 0.34 | | 2.32 | 2.08 | 2.58 | |

| | | | | | | | | |
|---|---|---|---|---|---|---|---|---|
| Leukaemia | 1.08 | 1.01 | 1.16 | | 0.38 | 0.35 | 0.41 | |
| Liver | 1.71 | 1.60 | 1.83 | | 0.39 | 0.36 | 0.42 | |
| Lung | 1.18 | 1.13 | 1.23 | | 0.88 | 0.84 | 0.92 | |
| Melanoma | 0.05 | 0.05 | 0.06 | | 4.14 | 3.92 | 4.37 | |
| Mesothelioma | 1.17 | 1.07 | 1.29 | | 0.70 | 0.63 | 0.78 | |
| Myeloma | 1.19 | 1.11 | 1.28 | | 0.61 | 0.57 | 0.66 | |
| NHL | 0.86 | 0.82 | 0.91 | | 0.86 | 0.82 | 0.90 | |
| Oesophagus | 0.54 | 0.50 | 0.57 | | 1.85 | 1.75 | 1.96 | |
| Oral | 0.20 | 0.18 | 0.22 | | 2.29 | 2.16 | 2.42 | |
| Other | 1.34 | 1.28 | 1.40 | | 0.55 | 0.52 | 0.57 | |
| Ovary | 1.08 | 1.01 | 1.15 | | 1.13 | 1.07 | 1.21 | |
| Pancreas | 1.78 | 1.69 | 1.88 | | 0.54 | 0.51 | 0.58 | |
| Prostate | 0.19 | 0.18 | 0.20 | | 1.94 | 1.86 | 2.02 | |
| Rectum | 0.34 | 0.32 | 0.37 | | 1.88 | 1.79 | 1.98 | |
| Renal | 0.69 | 0.65 | 0.74 | | 0.92 | 0.87 | 0.97 | |
| Stomach | 0.96 | 0.89 | 1.02 | | 1.02 | 0.96 | 1.09 | |
| Testis | 0.26 | 0.20 | 0.34 | | 3.56 | 3.08 | 4.12 | |
| Thyroid | 0.24 | 0.20 | 0.28 | | 0.72 | 0.65 | 0.79 | |
| Uterus | 0.22 | 0.20 | 0.24 | | 2.98 | 2.81 | 3.16 | |
| Learning disability (yes) | 2.65 | 2.26 | 3.11 | P<0.001 | 0.51 | 0.43 | 0.60 | P<0.001 |
| *Morbidity burden* | | | | P<0.001 | | | | P<0.001 |
| No morbidity burden | 1.00 | 1.00 | 1.00 | | 1.00 | 1.00 | 1.00 | |
| Low morbidity burden | 0.87 | 0.84 | 0.91 | | 0.95 | 0.93 | 0.97 | |
| Medium morbidity burden | 0.95 | 0.92 | 0.99 | | 0.91 | 0.89 | 0.94 | |
| High morbidity burden | 1.18 | 1.14 | 1.23 | | 0.75 | 0.73 | 0.77 | |
| Sex (Female) | 1.02 | 1.00 | 1.05 | 0.0397 | 0.97 | 0.95 | 0.99 | 0.0144 |
| *Age* | | | | P<0.001 | | | | P<0.001 |
| 40 to 44 years | 0.87 | 0.80 | 0.94 | | 0.79 | 0.74 | 0.84 | |
| 45 to 49 years | 0.92 | 0.86 | 0.98 | | 0.83 | 0.79 | 0.87 | |
| 50 to 54 years | 0.92 | 0.87 | 0.97 | | 0.84 | 0.80 | 0.87 | |
| 55 to 59 years | 0.94 | 0.90 | 0.99 | | 0.90 | 0.87 | 0.94 | |
| 60 to 64 years | 0.95 | 0.91 | 1.00 | | 0.96 | 0.93 | 0.99 | |
| 65 to 69 years | 1.00 | 1.00 | 1.00 | | 1.00 | 1.00 | 1.00 | |
| 70 to 74 years | 1.15 | 1.10 | 1.19 | | 1.01 | 0.98 | 1.04 | |
| 75 to 79 years | 1.32 | 1.27 | 1.37 | | 1.05 | 1.01 | 1.08 | |
| 80 to 84 years | 1.78 | 1.71 | 1.84 | | 0.91 | 0.88 | 0.94 | |
| 85 to 89 years | 2.67 | 2.56 | 2.79 | | 0.74 | 0.71 | 0.77 | |
| 90 years and older | 4.44 | 4.21 | 4.68 | | 0.49 | 0.46 | 0.51 | |
| *IMD* | | | | P<0.001 | | | | P<0.001 |
| 1 (least deprived | 1.00 | 1.00 | 1.00 | | 1.00 | 1.00 | 1.00 | |
| 2 | 1.09 | 1.05 | 1.12 | | 1.07 | 1.04 | 1.10 | |
| 3 | 1.16 | 1.12 | 1.20 | | 1.11 | 1.07 | 1.15 | |
| 4 | 1.28 | 1.24 | 1.33 | | 1.07 | 1.03 | 1.11 | |
| 5 (most deprived) | 1.42 | 1.37 | 1.47 | | 1.06 | 1.02 | 1.10 | |
| Smoking history | 1.02 | 1.00 | 1.05 | 0.0674 | 1.08 | 1.06 | 1.11 | P<0.001 |
| *Year of diagnosis* | | | | P<0.001 | | | | P<0.001 |
| 2012 | 1.00 | 1.00 | 1.00 | | 1.00 | 1.00 | 1.00 | |
| 2013 | 0.95 | 0.92 | 0.99 | | 1.02 | 0.99 | 1.05 | |
| 2014 | 0.97 | 0.93 | 1.00 | | 1.07 | 1.03 | 1.11 | |
| 2015 | 0.91 | 0.87 | 0.94 | | 1.15 | 1.11 | 1.19 | |
| 2016 | 0.87 | 0.84 | 0.91 | | 1.23 | 1.19 | 1.27 | |
| 2017 | 0.83 | 0.80 | 0.86 | | 1.28 | 1.24 | 1.33 | |
| 2018 | 0.84 | 0.81 | 0.87 | | 1.34 | 1.30 | 1.39 | |

**Table 4. Main effect only model associations between cancer stage, 30-day mortality, patient characteristics, cancer type and learning disability**

| | Advanced-stage cancer diagnosis | | | | 30-day mortality | | | |
|---|---|---|---|---|---|---|---|---|
| | Odds ratio | Lower 95% confidence interval | Upper 95% confidence interval | P | Odds ratio | Lower 95% confidence interval | Upper 95% confidence interval | P |
| *Cancer type* | | | | P<0.001 | | | | P<0.001 |
| Bladder | 0.24 | 0.23 | 0.26 | | 0.56 | 0.50 | 0.63 | |
| Brain | | | | | 1.62 | 1.43 | 1.83 | |
| Breast | 0.21 | 0.21 | 0.22 | | 0.29 | 0.26 | 0.32 | |
| Cervix | 0.46 | 0.40 | 0.53 | | 0.73 | 0.53 | 0.99 | |
| Colon | 1.00 | 1.00 | 1.00 | | 1.00 | 1.00 | 1.00 | |
| DCIS | 1.00 | 1.00 | 1.00 | | 0.02 | 0.00 | 0.06 | |
| HL | 1.04 | 0.91 | 1.20 | | 0.56 | 0.39 | 0.81 | |
| Larynx | 0.68 | 0.61 | 0.76 | | 0.53 | 0.41 | 0.68 | |
| Leukaemia | | | | | 1.56 | 1.42 | 1.71 | |
| Liver | 1.74 | 1.57 | 1.91 | | 2.98 | 2.74 | 3.25 | |
| Lung | 2.33 | 2.23 | 2.43 | | 2.32 | 2.19 | 2.46 | |
| Melanoma | 0.08 | 0.07 | 0.09 | | 0.07 | 0.05 | 0.09 | |
| Mesothelioma | 1.68 | 1.46 | 1.94 | | 1.13 | 0.98 | 1.31 | |
| Myeloma | 0.36 | 0.32 | 0.41 | | 0.77 | 0.68 | 0.87 | |
| NHL | 1.61 | 1.52 | 1.70 | | 0.97 | 0.89 | 1.06 | |
| Oesophagus | 2.15 | 2.01 | 2.30 | | 1.07 | 0.97 | 1.17 | |
| Oral | 2.07 | 1.93 | 2.22 | | 0.34 | 0.28 | 0.40 | |
| Other | 1.14 | 1.07 | 1.21 | | 3.49 | 3.28 | 3.71 | |
| Ovary | 1.32 | 1.24 | 1.41 | | 1.50 | 1.36 | 1.66 | |
| Pancreas | 2.79 | 2.61 | 2.99 | | 3.65 | 3.41 | 3.92 | |
| Prostate | 0.54 | 0.52 | 0.56 | | 0.17 | 0.16 | 0.19 | |
| Rectum | 1.00 | 0.95 | 1.05 | | 0.54 | 0.49 | 0.60 | |
| Renal | 0.67 | 0.63 | 0.71 | | 0.88 | 0.79 | 0.97 | |
| Stomach | 1.83 | 1.70 | 1.98 | | 1.54 | 1.39 | 1.69 | |
| Testis | 0.08 | 0.06 | 0.10 | | 0.28 | 0.13 | 0.61 | |
| Thyroid | 0.57 | 0.51 | 0.64 | | 0.61 | 0.47 | 0.78 | |
| Uterus | 0.20 | 0.18 | 0.21 | | 0.33 | 0.28 | 0.38 | |
| Learning disability | | | | 0.002 | | | | P<0.001 |
| (Yes) | 1.37 | 1.12 | 1.67 | | 3.77 | 3.10 | 4.59 | |
| *Morbidity burden* | | | | P<0.001 | | | | P<0.001 |
| No morbidity burden | 1.00 | 1.00 | 1.00 | | 1.00 | 1.00 | 1.00 | |
| Low morbidity burden | 0.83 | 0.80 | 0.85 | | 0.83 | 0.78 | 0.88 | |
| Medium morbidity burden | 0.80 | 0.78 | 0.83 | | 0.93 | 0.88 | 0.99 | |
| High morbidity burden | 0.71 | 0.69 | 0.74 | | 1.17 | 1.10 | 1.24 | |
| Sex (Female) | 0.87 | 0.85 | 0.90 | P<0.001 | 0.93 | 0.90 | 0.96 | P<0.001 |
| *Age* | | | | P<0.001 | | | | P<0.001 |
| 40 to 44 years | 0.64 | 0.60 | 0.69 | | 0.31 | 0.25 | 0.38 | |
| 45 to 49 years | 0.76 | 0.72 | 0.81 | | 0.43 | 0.38 | 0.50 | |
| 50 to 54 years | 0.85 | 0.81 | 0.89 | | 0.55 | 0.49 | 0.61 | |
| 55 to 59 years | 0.88 | 0.85 | 0.92 | | 0.65 | 0.60 | 0.71 | |
| 60 to 64 years | 0.95 | 0.91 | 0.98 | | 0.83 | 0.78 | 0.89 | |
| 65 to 69 years | 1.00 | 1.00 | 1.00 | | 1.00 | 1.00 | 1.00 | |
| 70 to 74 years | 1.04 | 1.01 | 1.08 | | 1.26 | 1.19 | 1.33 | |
| 75 to 79 years | 1.10 | 1.06 | 1.14 | | 1.49 | 1.41 | 1.58 | |
| 80 to 84 years | 1.20 | 1.15 | 1.25 | | 2.15 | 2.04 | 2.28 | |
| 85 to 89 years | 1.38 | 1.32 | 1.44 | | 3.23 | 3.05 | 3.43 | |
| 90 years and older | 1.53 | 1.42 | 1.63 | | 5.79 | 5.41 | 6.20 | |
| *IMD* | | | | P<0.001 | | | | P<0.001 |
| 1 (least deprived | 1.00 | 1.00 | 1.00 | | 1.00 | 1.00 | 1.00 | |
| 2 | 1.02 | 0.99 | 1.05 | | 1.12 | 1.07 | 1.18 | |
| 3 | 1.05 | 1.02 | 1.08 | | 1.18 | 1.13 | 1.24 | |
| 4 | 1.10 | 1.06 | 1.13 | | 1.29 | 1.23 | 1.35 | |

| | | | | | | | | |
|---|---|---|---|---|---|---|---|---|
| 5 (most deprived) | 1.13 | 1.09 | 1.16 | | 1.39 | 1.33 | 1.46 | |
| Smoking history | 1.05 | 1.03 | 1.07 | P<0.001 | 1.12 | 1.08 | 1.16 | P<0.001 |
| *Year of diagnosis* | | | | 0.066 | | | | P<0.001 |
| 2012 | 1.00 | 1.00 | 1.00 | | 1.00 | 1.00 | 1.00 | |
| 2013 | 0.99 | 0.96 | 1.03 | | 0.94 | 0.89 | 0.99 | |
| 2014 | 0.98 | 0.94 | 1.01 | | 0.83 | 0.79 | 0.88 | |
| 2015 | 0.98 | 0.94 | 1.01 | | 0.79 | 0.75 | 0.83 | |
| 2016 | 0.96 | 0.92 | 0.99 | | 0.72 | 0.69 | 0.76 | |
| 2017 | 0.97 | 0.94 | 1.01 | | 0.70 | 0.67 | 0.74 | |
| 2018 | 0.95 | 0.92 | 0.98 | | 0.70 | 0.66 | 0.74 | |

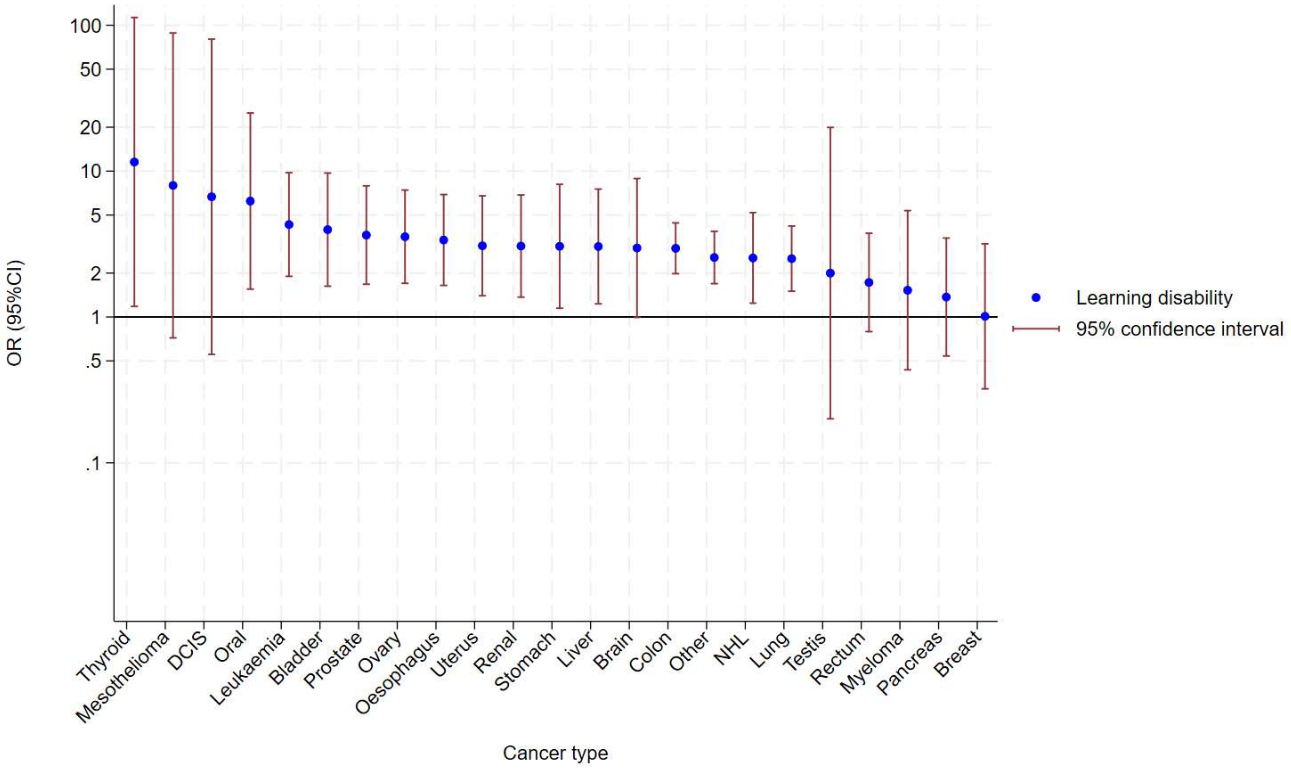


Figure 1. Associations between an emergency route to diagnosis and learning disability by cancer type

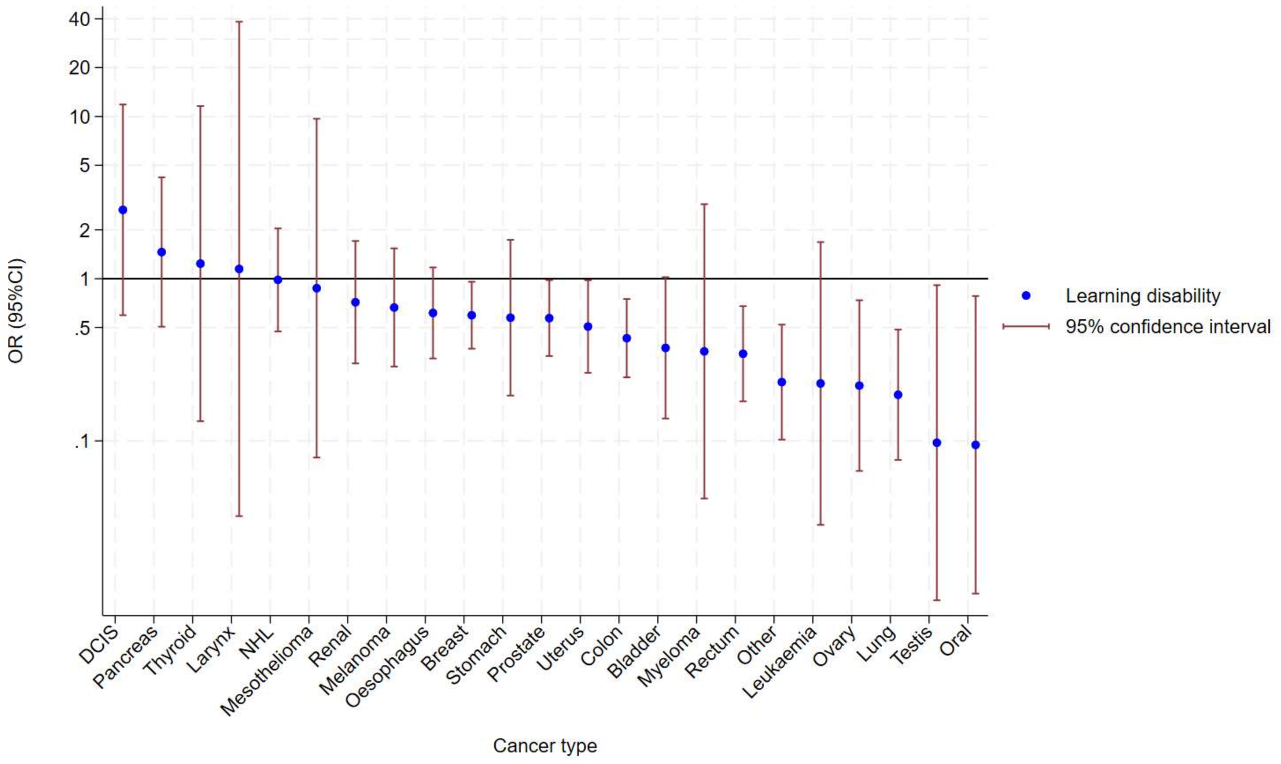


Figure 2. Associations between an urgent suspected cancer referral (two week wait) route to diagnosis and learning disability by cancer type

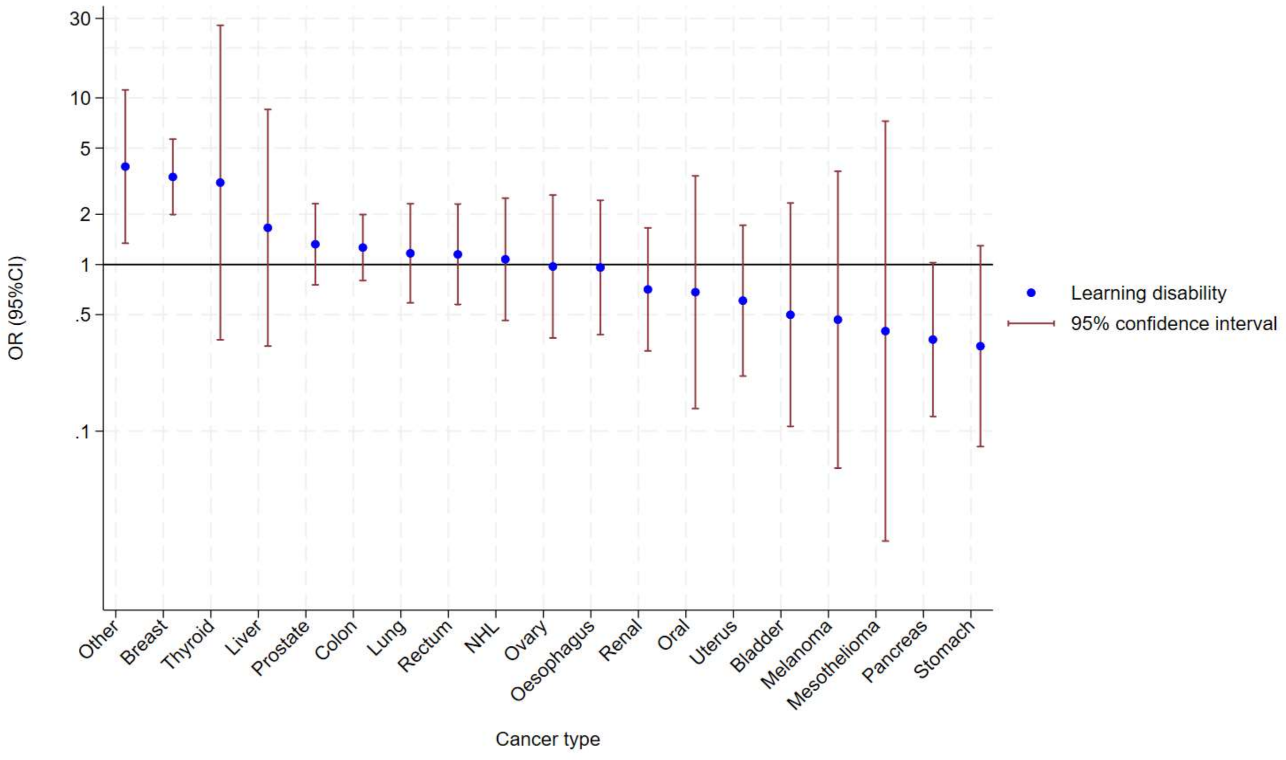


Figure 3. Associations between cancer stage and learning disability by cancer type

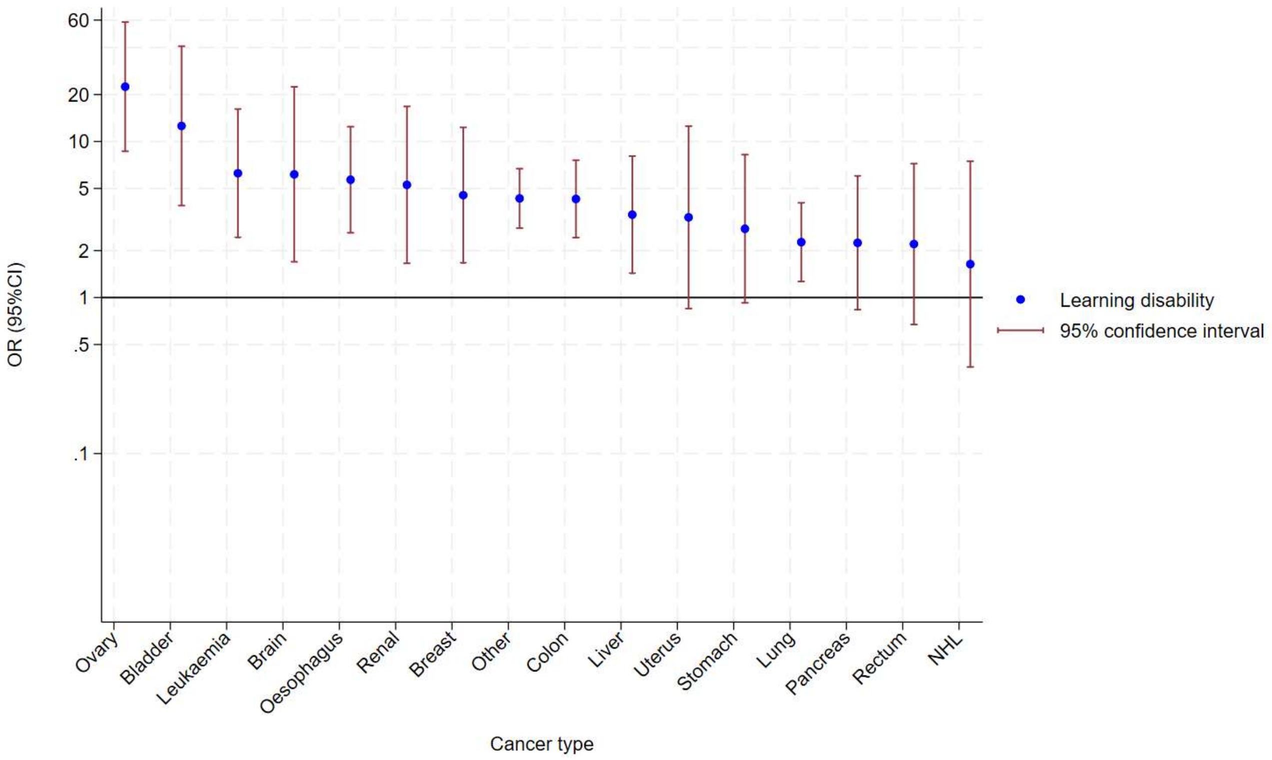


Figure 4. Associations between 30-day mortality and learning disability by cancer type
